# Congenital diaphragmatic eventration: Should we maintain surgical treatment? A retrospective multicentric cohort study

**DOI:** 10.1101/2024.01.25.24301768

**Authors:** Lymeymey Heng, Khalid Alzahrani, Naziha Khen-Dunlop, Nicoleta Panait, Erik Hervieux, Lucie Grynberg, Olivier Abbo, Frédéric Hameury, Frédéric Lavrand, Olivier Maillet, Aurore Haffreingue, Anne Lehn, Stephan De Napoli Cocci, Edouard Habonimana, Jean-Luc Michel, Louise Montalva, Quentin Ballouhey, Arnaud Fotso Kamdem, Jean-François Lecompte, Antoine Liné, Anna Poupalou, Pierre Maignan, Loren Deslandes, Guillaume Podevin, Françoise Schmitt

**Affiliations:** Paediatric Surgery Unit, Federation of paediatrics, University Hospital, Angers, France; Paediatric Surgery Department, Hôpital Necker-Sick Children, AP-HP, Paris, France; Paediatric Surgery Department, AP-HM, Marseille, France; Paediatric Surgery Department, Trousseau Hospital, AP-HP, Paris, France; Paediatric Surgery Department, University Hospital, Rouen, France; Paediatric Surgery Department, University Hospital, Toulouse, France; Paediatric Surgery Department, Hospices civils de Lyon, Bron, France; Paediatric Surgery Department, University Hospital, Bordeaux, France; Paediatric Surgery Department, University Hospital, Montpellier, France; Paediatric Surgery Department, University Hospital, Caen, France; Paediatric Surgery Department, Hôpital de Hautepierre, University Hospital, Strasbourg, France; Paediatric Surgery Department, University Hospital, Nantes, France; Paediatric Surgery Department, South Hospital, University Hospital, Rennes, France; Paediatric Surgery Department Nord, University Hospital -Réunion, Saint-Denis, France; Paediatric Surgery Department, Robert Debré Hospital, AP-HP, Paris, France; Paediatric Surgery Department, University Hospital, Limoges, France; Paediatric Surgery Department, University Hospital, Besançon, France; Paediatric Surgery Department, University Hospital-Lenval, Nice, France; Paediatric Surgery Department, University Hospital, Reims, France; Paediatric Surgery Department, HUDERF, Bruxelles, Belgique; Paediatric Surgery Department, University Hospital, Tours, France; Paediatric Surgery Department, University Hospital, Clermont-Ferrand, France

**Author notes:** **Corresponding author:** Françoise Schmitt, MD, PhD, Paediatric Surgery Department, University Hospital of Angers 4, Rue Larrey; 49933 ANGERS cedex 9, FRANCE, Mail. **Funding:** This research did not receive any specific grant from funding agencies in the public, commercial, or not-for-profit sectors. **Competing Interests** The authors declare that they have no competing interests.

**Keywords:** congenital diaphragmatic eventration, diaphragmatic plication, thoracoscopic plication, mini-invasive surgery, robot-assisted thoracoscopy

## Abstract

**Background:** Congenital diaphragmatic eventration (CDE) is an infrequent congenital pathology without consensus of treatment. This study assessed current care practices for this pathology in children in France.

**Methods:** Retrospective study on cohort data conducted in 22 paediatric surgery departments, including patients less than 16 years of age followed for CDE since 2010. Patients with surgical or conservative treatment were compared.

**Results:** 139 patients were included, with a median age of 8 [1 – 16] months. CDE occurred in boys in 68.3% and was right-sided in 66.7% of the cases. It was symptomatic in 65% of the cases, with a respiratory component for 97% of these patients. The primary indication for treatment, essentially depending on the clinical respiratory impairment and the level of the diaphragm, was surgery for diaphragmatic plication in 87 cases (62%) or clinical follow-up for the 52 others, 25 of whom were operated on secondarily. There were 32 early per- or postoperative complications (29%), and 8 recurrences of eventration (7%). With a median follow-up of 28 months, the median level of diaphragmatic dome fell from the 6th to the 9th back rib, and the rate of respiratory symptoms from 64% to 14%, in both surveyed and surgically treated patients.

**Conclusions:** CDE is mainly diagnosed in newborn or infant boys and right-sided. Diaphragmatic plication may be the best treatment in symptomatic patients with a dome level upper the 6^th^ posterior rib, but exposes them to a 29% complication rate and 7% of recurrence.

ClinicalTrials NCT04862494, April 28, 2021.

**What’s new?:** Boys are indeed more affected than girls (68.3%), but left-side eventration only happens in 33% of cases.

As compared to conservative management, surgery may be positively associated with diaphragmatic level improvement, but not with long term resolution of symptoms.

## Introduction

Congenital diaphragmatic eventration (CDE) is a rare defect in the central muscular portion of the diaphragm, caused by an abnormal migration of myoblast from the upper cervical somites during embryogenesis [1, 2]. Occurring in less than 0.05% of newborns, it causes paradoxical motion of the diaphragm as well as a lowering in the thoracic space for lung expansion, leading to impaired ventilation, from mild dyspnea to neonatal respiratory distress, asthma, recurrent infections, etc. When left-sided, it can lead to gastric ascension and plication causing impaired gastric emptying associated with dysphagia and vomiting [3]. In the literature, a surgical treatment, consisting in a diaphragmatic plication, has to be proposed to symptomatic patients [1, 4, 5], with immediate and long-term good results [6], but some authors advise surgery even in asymptomatic patients with a significant ascent of the diaphragmatic dome [4, 7] to preserve lung development and function in neonates.

The aims of the study were to (1) describe current practices regarding CDE management for CDE in children in France, and (2) compare outcomes of surgical and conservative treatment of CDE.

## Material and Methods

### 1. Population

We conducted a retrospective multicentric cohort study in all willing French and French speaking paediatric surgery departments. All patients diagnosed with CDE under the age of 16 between 2010-01-01 and 2021-08-31, were included in this study. Patients’ and parents’ consent to this inclusion was sought through an information letter addressed to the parents. First screening for eligible patients was completed thanks to French diagnosis and treatment classification. Exclusion criteria then were diaphragmatic hernia and acquired diaphragmatic eventration, defined as secondary to surgery, obstetrical trauma or oncologic aetiology.

Clinical factors recorded were: sex, age at diagnosis and treatment, associated non-causal pathologies, relevant associated pulmonary, digestive and orthopaedics symptoms, side of the eventration, results of medical imaging studies including thoracic radiographs, CT scan and/or magnetic resonance imaging (MRI). Late diagnosis was defined as a diagnosis beyond 12 months of age. Management data included the choice, reason and type of surgical or medical treatment, and if applicable, the occurrence of peroperative complications and early postoperative events. Postoperative and long-term survey data were clinical symptoms and level of the diaphragmatic dome on chest radiographs, complications with their severity level assessed through the Clavien-Dindo classification [8], need for additional hospital stays or re-operations.

Conservative treatment was defined as clinical and radiological follow-up. Failure of conservative treatment was defined as the need for diaphragmatic plication. Success of treatment was defined as the improvement or the resolution of symptoms.

### 2. Outcome measures

The primary outcome measure was the linear description of the cohort of paediatric patients, to expose current trends of care. Comparison between surgical and conservative treatments was a secondary outcome. A subgroup analysis of the symptomatic patients at baseline was performed too, with a comparative study between patients whose symptoms resolved or persisted.

### 3. Ethics

This study was performed in line with the principles of the Declaration of Helsinki. Approval was granted by the Ethics Committee of the University Hospital of Angers and is registered under the reference 2021-013. The associated registry has been approved by the French National Commission on Informatics and Liberty CNIL (Commission Nationale de l’Informatique et des Libertés, authorization ar21-001v0). The study has been registered in ClinicalTrials.gov under the identifier number NCT04862494 (April 28, 2021).

### 4. Statistical analysis

Statistical analysis was performed using IBM-SPSS 29.0.0.0 and GraphPad Prism 8.0.2 for Windows software. All tests were 2-sided and the statistical level for significance was set at p < 0.05. Patients’ characteristics were described as median with interquartile range or mean +/- standard deviation for continuous variables and as percentages for qualitative variables. Comparative analysis between groups of patients operated on or with conservative treatment was performed using the non parametric Mann-Whitney test for quantitative variables after assessing the absence of normality of the distribution with a Shapiro-Wilk normality test, and using the Fisher’s exact test for qualitative variables. Factors associated with occurrence to surgical treatment and efficiency of surgical treatment (vs conservative treatment) and with success (determined as resolution of symptoms) were determined using logistic regression. First, univariate analysis was used to select relevant variables for each of the above mentioned items, and variables with a p-value below 20% entered in the final model.

Multivariate analysis was carried out using a backward stepwise likelihood-ratio test and data were expressed as odds-ratio (OR) with 95% confidence intervals.

## Results

### 1. Description of the patients

Among the 22 paediatric surgery departments which took part in the study, there were 175 patients with a diagnosis of diaphragmatic eventration. Thirty six of them (20.6%) were excluded from further analysis because of their acquired mechanism, as detailed on the flowchart (**Fig. 1**).

**Fig. 1.**
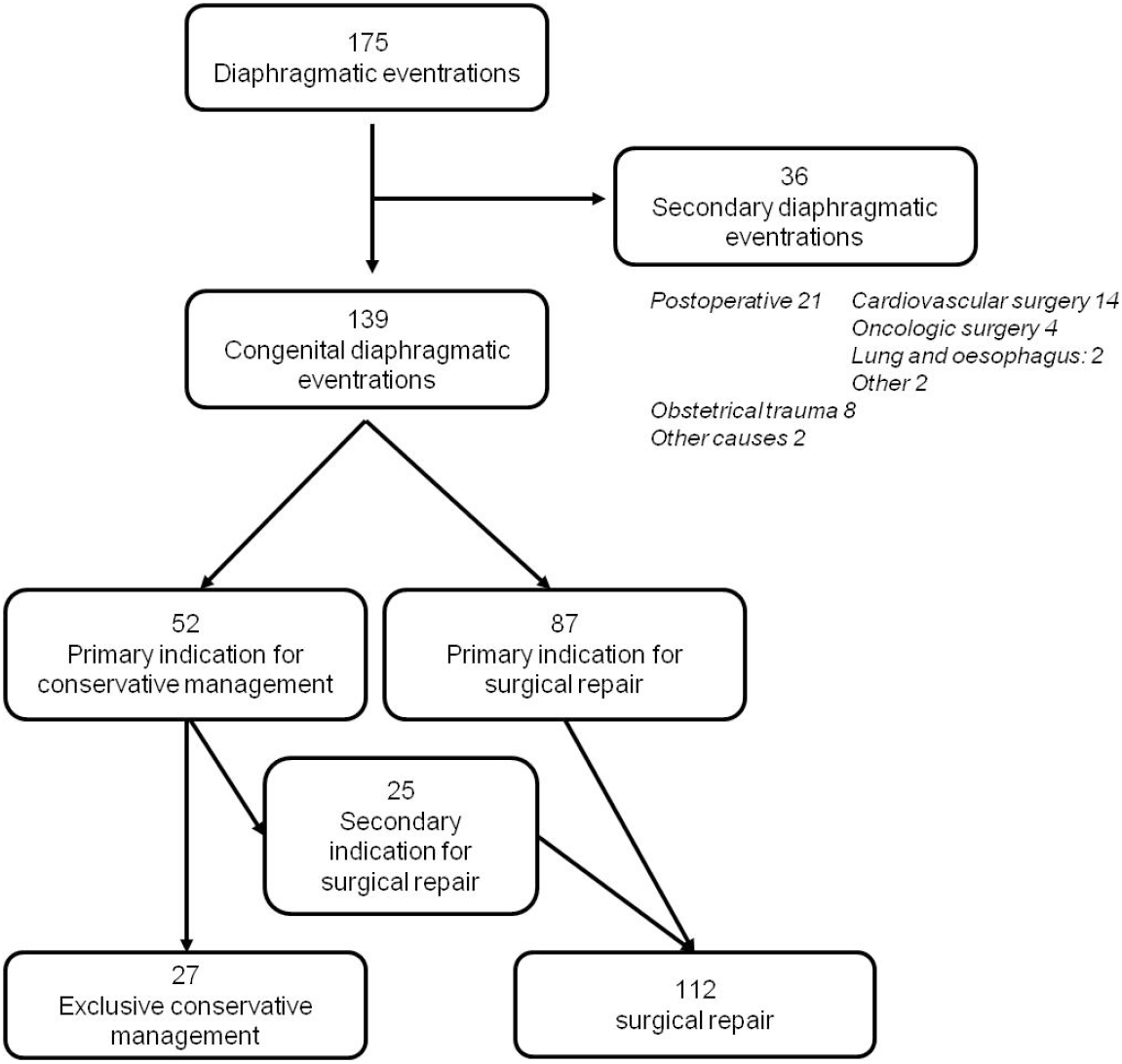
Flowchart of the cohort of paediatric patients with diaphragmatic eventration

The study population included 139 patients with CDE, composed of 68.3% boys, with a mean gestational age at birth of 37.2 +/- 3.8 weeks. Diaphragmatic eventration was left-sided in 33.1%, with only one case of bilateral CDE. Associated pathologies were present in 48 (34.5%) patients, including 12 syndromic or genetic forms, 10 cardiovascular pathologies, 13 orthopaedic abnormalities, 7 digestive afflictions (including 3 cases of inguinal hernia), 6 uro-genital malformations and 5 associated broncho-pulmonary abnormalities. Median age at diagnosis was 8 [1 – 16] months], with 39 patients (28.1%) having a late diagnosis (>12 months). Ninety patients (64.7%) presented with symptoms, mainly respiratory (n = 87), with 33 (37.9%) of them presenting obstructive signs (asthma, bronchial hyper-reactivity, bronchiolitis), 31 (35.6%) with neonatal respiratory distress or later dyspnea, and 20 (23.0%) with recurrent pulmonary infections. Orthopaedic abnormalities (scoliosis or chest asymmetry) occurred in 9 (6.5%) patients and gastro-intestinal symptoms, in 17 (11.2%). Among the latter, clinical presentation did not differ according to the CDE side, with 8 patients presenting with gastro-oesophageal reflux (47%), 5 (29%) with failure to thrive and 4 (23.5%) with anorexia, dysphagia or vomiting/upper occlusive signs, but appeared to be more present in left-sided CDE (10/46, 21.7%) than in right-sided ones (7/94, 7.4%, p = 0.025).

All patients had chest radiographs, showing a dome at a median level of the 6^th^ posterior rib [5-7], and 84 (60.4%) of them completed the diagnosis by CT-scan or MRI, on which 50 (59.5%) did not have associated pulmonary abnormalities. No correlation could be found between the presence of symptoms and the level of the dome or CT-scan pulmonary abnormalities.

Initial treatment choice was conservative treatment for 52 (37.4%) patients, with a median age of 2 [0 – 10] month at diagnosis. With a median follow-up of 9 [4 – 36] months, this conservative treatment appeared to be sufficient in 27 patients (52%). However, it failed in 25 (48%), that eventually underwent surgery at a median age of 13.0 [3.5 – 21.5] months, after a median delay of 90 [16 - 330] days. The other 87 (62.6%) patients had a primary indication for surgery. After a median 23.5 [7.0 - 57.3] months of survey of the whole cohort of patients, only 30 (21.6%) patients presented with symptoms, and 22 of them (19.6%) had experienced surgical complications. On the last chest radiography, median level of the dome was set at the 8^th^ intercostal space [rib 8 - 9], with an improvement of this level for 77 (81.9%) patients and stabilization for 14 (14.9%) others.

Among the patients presenting with symptoms at baseline, 22 (25.3%) had persistent symptomatology at the end of survey, regardless of their treatment group. On univariate analysis, there was no difference between them and resolved patients (n = 65) in terms of baseline characteristics, treatment options and outcomes, except for the sex ratio (13.6% vs 41.5% girls, p = 0.02) and the absence of surgical complication (27.3% vs 0%, p<0.0001). After logistic regression, only the female sex appeared to be predictive for symptom resolution (OR = 10.5 [1.3 – 88.8], p = 0.03).

### 2. Comparison of operated and non operated patients’ outcomes

#### Baseline characteristics

A total of 112 patients (80.5%) eventually underwent surgical treatment, whereas 27 (19.5%) were managed using conservative treatment (CoT group). Both groups were similar in terms of age at diagnosis, associated comorbidities and side of the eventration (**Table 1**). However, patients that underwent surgical treatment were 1.7 times more likely to be symptomatic (72.3% vs 40.7%, p = 0.003), had a higher dome elevation (6 [5 – 7] vs 7 [6 – 8], p < 0.0001) and were more likely to have pulmonary abnormalities on CT-scan (57.8% vs 20.0%, p <0.0001). To note, none of these factors appeared to be associated with the choice of treatment on multivariate analysis.

**Table 1:**
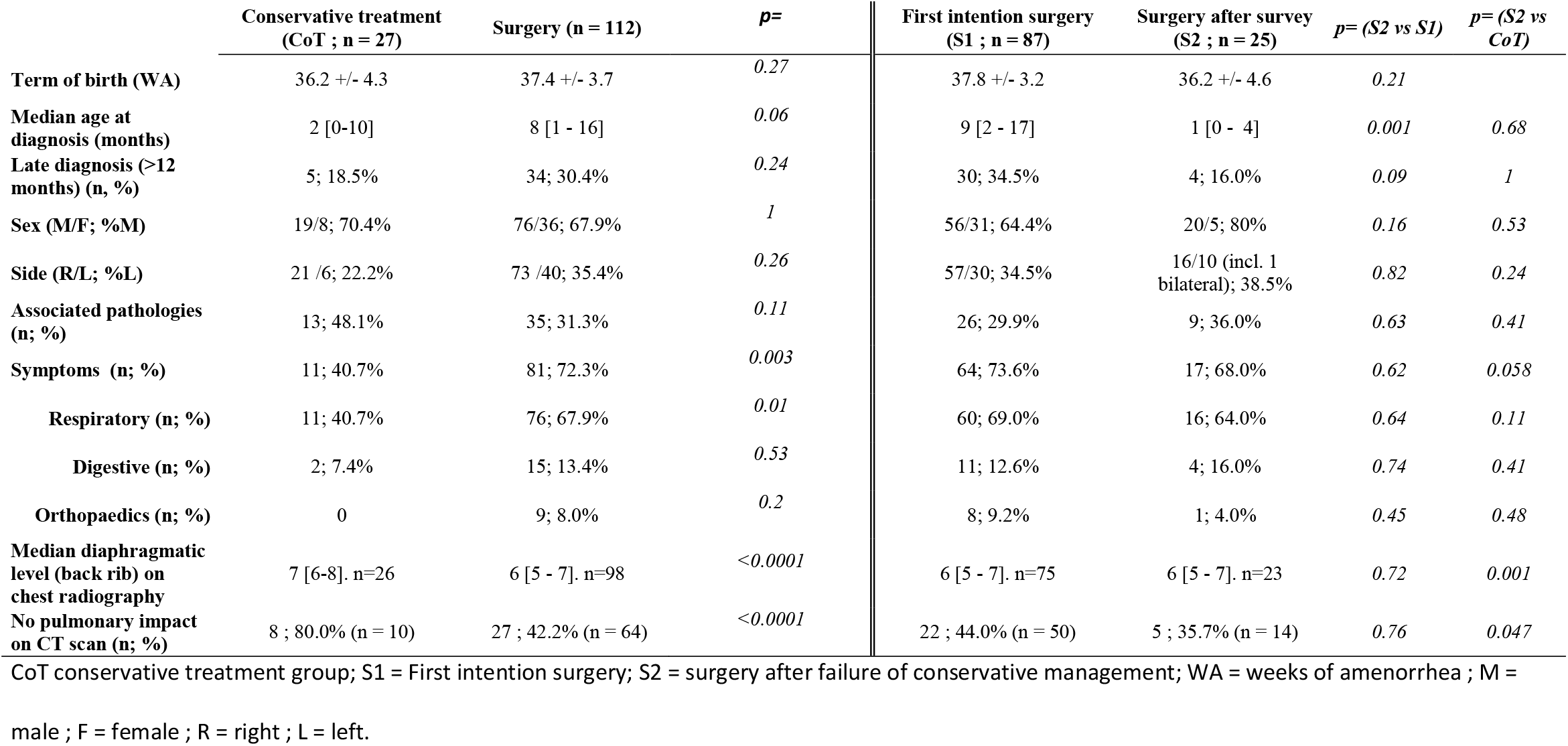
Univariate analysis of baseline characteristics of the patients according to their treatment.

A subgroup analysis of the patients operated on as first intention (S1 group, n = 87) or after failure of conservative treatment (S2 group, n = 25) showed no difference in terms of symptoms frequency, dome level or pulmonary abnormalities on CT-scan (**Table 1**). Median age at diagnosis in the S2 group (1 month [0 – 4]) was equivalent to the one of the CoT group (2 months [0 – 10]) and lower than in the S1 group (9 months [2 – 17], p=0.001). Nevertheless, in the S2 group, median duration of conservative treatment before surgery was 90 [16 – 330] days, resulting in similar age at surgery in the S1 and S2 groups (11.5 [5.8 – 21.1] vs 13.0 [3.5 – 21.5] months, p= 0.9).

#### Treatments’ outcomes

Among patients managed conservatively, none required further hospital stay due to diaphragmatic eventration until the end of follow-up (median 9 months [4 - 36]). Clinical evaluation showed persistent symptoms in 5 (18.5%) patients, all with respiratory events (asthma 3, pulmonary infections 2), and one with an associated thoracic asymmetry. On chest radiographs, the level of the dome was improved in 44.4% of the cases, stable in 27.8% and worsened in 11.1% (**Table2**).

**Table 2:**
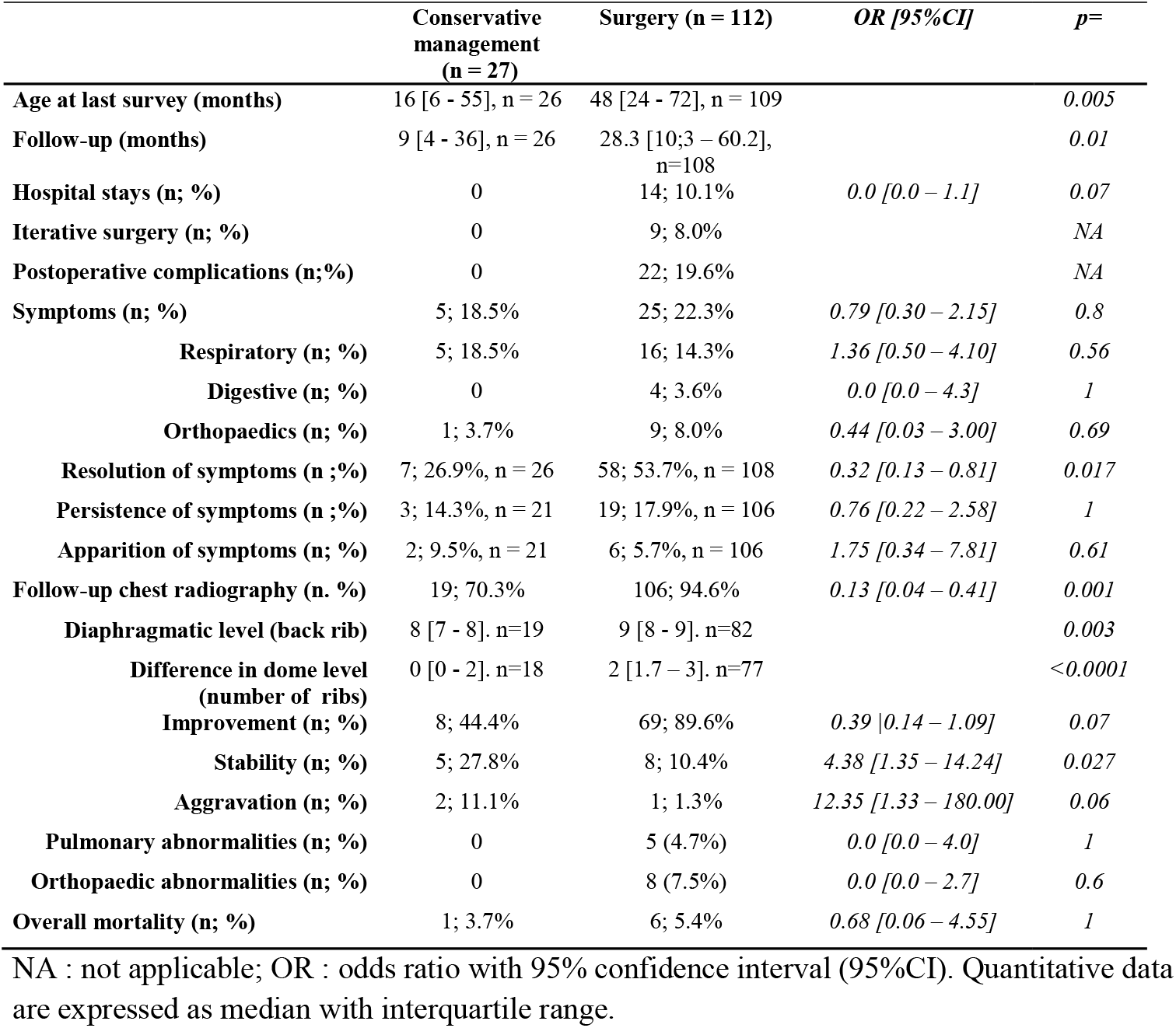
comparative outcomes of congenital diaphragmatic eventrations.

Surgery was performed at a median age of 12 [5 – 21] months. The surgical approach was abdominal for 28 (25%) patients (12 laparotomies and 16 laparoscopies) and thoracic in the remaining 84 (75%) others (thoracotomy 15, thoracoscopy 64 and robot-assisted thoracoscopy 5). After surgery, median duration for intravenous analgesia was 48 [12 – 72] hours, and median hospital stay 4 [3 – 7] days.

Perioperative complications occurred in 10 patients (8.9%), with 3 hepatic bleeding, 3 pulmonary or pleural breaks, two tracheal tube-related issues and one conversion from thoracoscopy to thoracotomy. There were 22 (19.6%) early postoperative complications, ten out of them being infections, 7 pneumothorax and one chylothorax, 4 atelectasis, 2 relapses of eventration. The repartition of gravity levels of postoperative complications was as follow: 10 Clavien-Dindo grade 1(45.5%); 5 grade 2 (22.7%), one of each for grades 3a and 3b (4.5%), and 3 grade 4a (13.5%).

In patients that were symptomatic, surgery allowed immediate resolution of respiratory symptoms in 88.3% (n=59/70). However, surgery was less effective in neonates, with only 53.8% (n = 7/13) of resolution of neonatal respiratory distress, as compared to 91% (n = 52/57) in symptomatic cases beyond the neonatal period (p = 0.003).

With a median postoperative follow-up of 28 months [10 - 60], 9 (8.0%) patients required surgical reintervention: for redo-plication for DE recurrence in 5 cases (4.5%), for diaphragmatic hernia or thoracic evisceration repair in 3 cases (2.7%), and for hiatal hernia repair associated to fundoplication in the last case. The median delay between initial surgery and reintervention was 6 [1 – 16] months. Twenty five patients (22.3%) remained symptomatic, with respiratory symptoms for 16 (14.3%) of them, including infections for 9 (8.0%), asthma for 6 (5.4%) and a restrictive syndrome for the remaining patient. Four patients presented with GERD (3.6%) and nine (8.0%) with orthopaedic disorders, including 6 scoliosis (4.3%) and 4 chest wall deformities (2.9%). On chest radiographs, the median level of the operated dome was at the 9^th^ rib [8 – 9], with an overall improvement of 2 costal levels [1.7 – 3.0], which was better than in the CoT group (0 [0 -2], p = 0.005). Pre and post-interventions comparison (**Table 2**) found some radiographic improvement in up to 69 (89.6%) cases, stability in 8 (10.4%) cases and worsening in 1 (1.3%) case; these proportions significantly differed from patients that underwent conservative treatment. Multivariate analysis of treatments outcomes confirmed that surgery improved the level of the dome (OR = 11.5, 95%CI [3.2 – 41.7], p<0.0001), but had no impact on the long term resolution of symptoms (OR = 2.1, 95%CI [0.6 – 7.4], p = 0.23).

## Discussion

With 139 patients over ten years, this study is one of the largest series currently published concerning CDE in children, first making it possible to specify its demographic characteristics. Hence, it has for long been advocated that CDE occurred in boys in almost 80% of the cases and was left-sided in the same proportions [9]. Interestingly, while there was indeed a majority of boys (68.3%) in our series, CDE only concerned the left dome in 46 cases (33.1%). As reported by Le Pimpec-Barthes *et al*. in 2010 [6], previous data were based on less than 150 patients cumulated from about ten studies. The integration of the data of this study with those of the publications made in the last twenty years [4, 5, 7, 10–12] now allows for a re-estimation of the rate of CDE in boys at 69.0% (347 out of 503 patients) and of left-sided at only 30.4% of the cases (173 / 570 patients).

As expected, most of the CDE were discovered during the first months of life, and a majority of the patients presented with symptoms, mostly in the respiratory tract. Among them, no higher representativeness of infectious signs, or obstructive or restrictive syndromes was found. Gastro-intestinal symptoms only occurred in 11% of the cases, and were more present when CDE was left-sided. These findings seem to be concordant with the literature [7, 9].

While the diagnosis and the level of the dome were assessed by chest radiographs in all patients, only 60.4% of them had additional imaging, which showed lung compression signs in 59% of the cases. No tumoral or anatomical cause to phrenic nerve compression was identified, which was a reason given by Le Pimpec-Barthes *et al*. [6] for the systematic performance of complementary examinations in late-onset forms. In this series, no correlation could be found between CT-scan pulmonary abnormalities and respiratory symptoms, indication for surgical treatment or subsequent clinical improvement. No functional imaging, such as diaphragm fluoroscopy or dynamic US or MRI, was reported in this series. Indeed, in showing the paradoxical motion of the diaphragm and its potential repercussion on lung expansion [13, 14], these exams may improve surgical indication and efficiency, but these hypotheses remain to be tested.

Surgical treatment has long been indicated for symptomatic patients [6, 9], and conservative management for asymptomatic or paucisymptomatic patients [15], even though some authors once advocated for systematic diaphragmatic plication to prevent lung growth restriction in neonates [4]. More recently, Wu *et al*. [5] have listed the respiratory symptoms relevant for surgery, and Zhao *et al*. [7] have proposed the subsequent criteria for surgical treatment: diaphragmatic elevation ≥ 3 intercostals spaces, presence of symptoms attributable to CDE, aggravation of the eventration during clinical follow-up. In this study, it appeared that criteria to surgery in our centres were the presence of symptoms and/or an elevation of the diaphragm ≥ 6^th^ posterior rib, equivalent to Zhao’s one.

After surgery, 88.3% of the patients experienced immediate resolution of symptoms, at the cost of a median 48 hours of intravenous analgesia, 4 days’ hospital stay, and a 28.6% rate of per- and postoperative complications, being graded serious (grades 3 – 4 of the Clavien-Dindo classification) in one third of the cases. When more specifically looking for neonatal respiratory distress, only 53.8% of the patients showed disappearance of symptoms, but our study could not qualify partial respiratory improvement, which may be sufficient for surgical success, as their underlying respiratory condition may be impacted by other factors, such as prematurity, surfactant abnormalities, etc.

With more than 2 years of follow-up, 79% of the patients appeared to be clinically resolved, whatever their treatment option, while chest radiographs showed twice the improvement of the diaphragmatic level after surgery than with conservative management (44.4% vs 89.6%, OR = 11.5). On the whole, most series describe prompt and durable improvement or resolution of symptoms after surgery in 91 to 100% of the cases [4, 5, 7, 10, 14], with 0 to 7% of incomplete repair or eventration recurrence [14, 16, 17], which is comparable to our 7.1% recurrence rate. Another explanation for recurrence could be the difficulty to sometimes differentiate CDE from congenital diaphragmatic hernia with a hernia sac, even during surgery, the treatment of which consists of the resection of the sac and hernia repair rather than diaphragmatic plication [18].

Pulmonary function tests (PFTs) were not available for our patients, due to the fact that they were too young for this exam. Other studies in older paediatric patients or adults have shown that DE induced a decrease of 20-30% of total lung capacity [5], and that there was a postoperative recovery of 10-20% of the forced vital capacity as well as the Tiffeneau-Pinelli index [19–22]. Hence, long-term survey of CDE patients including PFTs may be interesting in assessing efficiency of surgical treatment or for the precocious detection of infra-clinical restrictive symptoms for surveyed patients.

The main weakness of our study is its retrospective and multicentric design, exposing it to selection biases such as over-missing non-operated patients or overestimating the number of symptomatic patients, the others often being lost to follow-up. Nevertheless, the rarity of CDE make any prospective monocentric study unrealistic, while this cohort observational study presents an honest reflection of current care practices all over the national territory. As well, association analysis has to be taken as indicative as the small size of comparison groups doesn’t always confer sufficient statistical power to definitely conclude. To objectively assess treatment efficiency, ideally systematic PFTs would have been performed, but our patients were mostly too young to have undergone this exam, and so this element could only be implemented in a long term survey in the future. An alternative could be the systematic use of clinical scores for dyspnoea in children. Nevertheless, this study remains one of the largest series of CDE paediatric patients yet published, and allows for a number of conclusions that may help to improve further practices.

## Conclusion

Congenital diaphragmatic eventration is a rare condition that may impair pulmonary or digestive function. Often diagnosed during the first weeks of life, it may require surgical treatment, but one should be cautious, with regards to the 28% rate of complications and 7% recurrence rate, and knowing that this clinical condition may spontaneously resolve in up to 27% of cases. Indeed, apart from the cases of acute respiratory distress or upper digestive tract occlusion where surgery appears to be essential, surgery has been more strongly associated here with diaphragmatic level improvement on chest radiograph than with symptom resolution.

## Data Availability

All data produced in the present study are available upon reasonable request to the authors

## Abbreviations

CDE: congenital diaphragmatic eventration
CNIL: Commission Nationale de l’Informatique et des Libertés
CT: Computed Tomography
DDL: diaphragmatic dome level
ICS: intercostals space
MRI: magnetic resonance imaging
NA: not applicable
OR: odds ratio
PFT: pulmonary function test
ROC: Receiver Operating Characteristics

## Acknowledgements

To Mr Ali Gillies from The Lanuage Room (Lyon, France), who performed the English proofreading of the manuscript.

## References

1. Gutt CN, Grabensee R (2015) Diaphragm Plication and Repair. In: Dienemann HC, Hoffmann H, Detterbeck FC (eds) Chest Surgery. Springer Berlin Heidelberg, Berlin, Heidelberg, pp 483–494

2. Patrini D, Panagiotopoulos N, Bedetti B, et al (2017) Diaphragmatic plication for eventration or paralysis. Shanghai Chest 1:25–25. 10.21037/shc.2017.08.01

3. Favre J-P, Favoulet P, Cheynel N, Benoit L (2005) Traitement chirurgical des éventrations diaphragmatiques. EMC - Chirurgie 2:235–241. 10.1016/j.emcchi.2005.03.001

4. Yazici M, Karaca I, Arikan A, et al (2003) Congenital Eventration of the Diaphragm in Children: 25 Years’ Experience in Three Pediatric Surgery Centers. Eur J Pediatr Surg 13:298–301. 10.1055/s-2003-43573

5. Wu S, Zang N, Zhu J, et al (2015) Congenital diaphragmatic eventration in children: 12years’ experience with 177 cases in a single institution. Journal of Pediatric Surgery 50:1088–1092. 10.1016/j.jpedsurg.2014.09.055

6. Le Pimpec-Barthes F, Brian E, Vlas C, et al (2010) Le traitement chirurgical des éventrations et paralysies diaphragmatiques. Revue des Maladies Respiratoires 27:565–578. 10.1016/j.rmr.2010.01.015

7. Zhao S, Pan Z, Li Y, et al (2020) Surgical treatment of 125 cases of congenital diaphragmatic eventration in a single institution. BMC Surg 20:270. 10.1186/s12893-020-00928-z

8. Dindo D, Demartines N, Clavien P-A (2004) Classification of surgical complications: a new proposal with evaluation in a cohort of 6336 patients and results of a survey. Ann Surg 240:205–213. 10.1097/01.sla.0000133083.54934.ae

9. Ortega Deballon P, Facy O, Cheynel N, Rat P (2012) Traitement chirurgical des éventrations diaphragmatiques. EMC - Techniques chirurgicales - Appareil digestif 7:1–6. 10.1016/S0246-0424(12)57344-9

10. Becmeur F, Talon I, Schaarschmidt K, et al (2005) Thoracoscopic diaphragmatic eventration repair in children: about 10 cases. Journal of Pediatric Surgery 40:1712–1715. 10.1016/j.jpedsurg.2005.07.008

11. Ghribi A, Bouden A, Braiki M, et al (2015) Diaphragmatic eventration in children. Tunis Med 93:76–78

12. Cai Y, Wu Y, Wu Z, et al (2021) Comparative Study of Thoracoscopic and Modified Small Incision Repair for Congenital Diaphragmatic Eventration in Children. J Laparoendosc Adv Surg Tech A 31:1079–1083. 10.1089/lap.2021.0110

13. Nason LK, Walker CM, McNeeley MF, et al (2012) Imaging of the Diaphragm: Anatomy and Function. RadioGraphics 32:E51–E70. 10.1148/rg.322115127

14. Bawazir OA, Banaja AM (2020) Thoracoscopic repair of diaphragmatic eventration in children: a comparison of two repair techniques. Journal of Pediatric Surgery 55:1152–1156. 10.1016/j.jpedsurg.2019.11.019

15. Puri P, Nakazawa N (2009) Congenital Diaphragmatic Hernia. In: Puri P, Höllwarth M (eds) Pediatric Surgery. Springer Berlin Heidelberg, Berlin, Heidelberg, pp 307–313

16. Fujishiro J, Ishimaru T, Sugiyama M, et al (2016) Minimally invasive surgery for diaphragmatic diseases in neonates and infants. Surg Today 46:757–763. 10.1007/s00595-015-1222-3

17. Borruto FA, Ferreira CG, Kaselas C, et al (2014) Thoracoscopic treatment of congenital diaphragmatic eventration in children: lessons learned after 15 years of experience. Eur J Pediatr Surg 24:328–331. 10.1055/s-0033-1349054

18. Heiwegen K, van Heijst AF, Daniels-Scharbatke H, et al (2020) Congenital diaphragmatic eventration and hernia sac compared to CDH with true defects: a retrospective cohort study. Eur J Pediatr 179:855–863. 10.1007/s00431-020-03576-w

19. Versteegh MIM, Braun J, Voigt PG, et al (2007) Diaphragm plication in adult patients with diaphragm paralysis leads to long-term improvement of pulmonary function and level of dyspnea. Eur J Cardiothorac Surg 32:449–456. 10.1016/j.ejcts.2007.05.031

20. Freeman RK, Van Woerkom J, Vyverberg A, Ascioti AJ (2009) Long-term follow-up of the functional and physiologic results of diaphragm plication in adults with unilateral diaphragm paralysis. Ann Thorac Surg 88:1112–1117. 10.1016/j.athoracsur.2009.05.027

21. Bin Asaf B, Kodaganur Gopinath S, Kumar A, et al (2021) Robotic diaphragmatic plication for eventration: A retrospective analysis of efficacy, safety, and feasibility. Asian J Endosc Surg 14:70–76. 10.1111/ases.12833

22. Ribet M, Linder JL (1992) Plication of the diaphragm for unilateral eventration or paralysis. Eur J Cardiothorac Surg 6:357–360. 10.1016/1010-7940(92)90172-t

